# Intermediate-dose anticoagulation, aspirin, and in-hospital mortality in COVID-19: a propensity score-matched analysis

**DOI:** 10.1101/2021.01.12.21249577

**Authors:** Matthew L. Meizlish, George Goshua, Yiwen Liu, Rebecca Fine, Kejal Amin, Eric Chang, Nicholas DeFilippo, Craig Keating, Yuxin Liu, Michael Mankbadi, Dayna McManus, Stephen Wang, Christina Price, Robert D. Bona, Cassius Iyad Ochoa Chaar, Hyung J. Chun, Alexander B. Pine, Henry M. Rinder, Jonathan Siner, Donna S. Neuberg, Kent A. Owusu, Alfred Ian Lee

**Affiliations:** Yale School of Medicine, New Haven, CT; Section of Hematology, Department of Medicine, Yale School of Medicine, New Haven, CT; Dana-Farber Cancer Institute, Boston, MA; Department of Medicine, Yale School of Medicine, New Haven, CT; Department of Pharmacy, Yale-New Haven Hospital, New Haven, CT; School of Pharmacy, University of Connecticut, Storrs, CT; Joint Data Analytics Team, Yale New Haven Hospital, New Haven, CT; Section of Allergy and Immunology, Department of Medicine, Yale School of Medicine, New Haven, CT; Section of Vascular Surgery, Department of Surgery, Yale School of Medicine, New Haven, CT; Section of Cardiology, Department of Medicine, Yale School of Medicine, New Haven, CT; Department of Laboratory Medicine, Yale School of Medicine, New Haven, CT; Section of Pulmonary and Sleep Medicine, Department of Medicine, Yale School of Medicine, New Haven, CT; Clinical Redesign, Yale New Haven Health, New Haven, CT

**Author notes:** Contributed equally as first authors (Matthew L. Meizlish, George Goshua, Yiwen Liu, Rebecca Fine). Contributed equally as senior authors (Donna S. Neuberg, Kent A. Owusu, Alfred Ian Lee). Corresponding authors: Alfred Ian Lee, M.D., PhD.; Kent Owusu, Pharm.D.; Donna S. Neuberg, Sc.D. Funding information: This work was supported by a gift donation from Jack Levin and a separate anonymous donation to the Benign Hematology program at Yale, the DeLuca Foundation to fund hematology research at Yale, and the National Institutes of Health (grant HL142818 to H.J.C., and GM136651 and HL139116 to M.L.M.). Notation of prior abstract presentation: This work was presented at an Oral Session at the 62^nd^Annual Meeting of the American Society of Hematology in December 2020 and was highlighted afterwards at a “Best of ASH” session by the Hemostasis and Thrombosis Research Society.

## Abstract

**Background:** Thrombotic complications occur at high rates in hospitalized patients with COVID-19, yet the impact of intensive antithrombotic therapy on mortality is uncertain.

**Research Question:** How does in-hospital mortality compare with intermediate-versus prophylactic-dose anticoagulation, and separately with in-hospital aspirin versus no antiplatelet therapy, in treatment of COVID-19?

**Study Design and Methods:** Using data from 2785 hospitalized adult COVID-19 patients, we established two separate, nested cohorts of patients (1) who received intermediate- or prophylactic-dose anticoagulation (“anticoagulation cohort”, N = 1624), or (2) who were not on home antiplatelet therapy and received either in-hospital aspirin or no antiplatelet therapy (“aspirin cohort”, N = 1956). Propensity score matching utilizing various markers of illness severity and other patient-specific covariates yielded treatment groups with well-balanced covariates in each cohort. The primary outcome was cumulative incidence of in-hospital death.

**Results:** Among propensity score-matched patients in the anticoagulation cohort (N = 382), in a multivariable regression model, intermediate-compared to prophylactic-dose anticoagulation was associated with a significantly lower cumulative incidence of in-hospital death (hazard ratio 0.518 [0.308-0.872]). Among propensity-score matched patients in the aspirin cohort (N = 638), in a multivariable regression model, in-hospital aspirin compared to no antiplatelet therapy was associated with a significantly lower cumulative incidence of in-hospital death (hazard ratio 0.522 [0.336-0.812]).

**Interpretation:** In this propensity score-matched, observational study of COVID-19, intermediate-dose anticoagulation and aspirin were each associated with a lower cumulative incidence of in-hospital death.

**Summary conflict of interest statements:** No conflict of interest exists for any author on this manuscript.

## INTRODUCTION

Thrombosis is among the most devastating complications of COVID-19. In multiple studies, venous thromboembolism (VTE), arterial thrombosis, and microvascular thrombosis have all been described.^1-6^ High VTE rates have been reported in critically ill COVID-19 patients despite the use of prophylactic anticoagulation.^1,6,7^ The development of pulmonary microvascular thrombosis may be central to the pathogenesis of COVID-19 in the lungs.^5^ An elevated D-dimer, a breakdown product of fibrin clots, is one of the strongest predictors of mortality from COVID-19.^8,9^

A common global practice has been to administer escalated intensities of antithrombotic therapy beyond standard prophylactic-dose anticoagulation in hospitalized COVID-19 patients.^10-12^ To date, there has been little evidence to support this practice.^13,14^ Some retrospective studies have observed lower mortality rates with therapeutic-dose anticoagulation compared to either prophylactic-dose anticoagulation or no anticoagulation, while others comparing therapeutic- and prophylactic-dose anticoagulation have found no mortality difference.^15-22^ Recent findings from the ACTIV randomized controlled trial suggested futility of therapeutic-compared to prophylactic-dose anticoagulation in critically ill COVID-19 patients, leading to a pausing of enrollment of critically ill patients in that trial. To date, however, no large-scale study has compared the effects of intermediate-versus prophylactic-dose anticoagulation. Some investigators have also proposed a potential role for aspirin and other antiplatelet therapies in light of the high burden of microvascular thrombosis and emerging models of immunothrombosis in COVID-19.^5,23,24^ One retrospective study reported improved outcomes with aspirin therapy but did not account for disease severity between treatment groups, making its conclusions difficult to interpret.^25^

A major limitation in retrospective studies is bias in the likelihood of patients to receive the treatments being studied. In unadjusted observational studies, disease severity is a confounding factor affecting treatment decisions and outcomes, often precluding accurate analysis of potential treatment effects. To address this, propensity score matching for disease severity and other variables has been utilized in some observational studies, leading to findings compatible with those obtained from randomized controlled trials.^26,27^ The use of propensity score matching in a few landmark observational studies in COVID-19 has yielded key insights about potential treatment effects by enabling treatment groups with balanced covariates to be reliably compared.^28,29^

We sought to examine the impact of intermediate-dose anticoagulation and aspirin on in-hospital mortality in COVID-19. To account for variations in treatment, we utilized propensity score matching and multivariable regression analysis incorporating markers of disease severity and other clinical covariates. One scoring system for assessing disease severity in use at many hospitals is the Rothman Index (RI), a composite score of 26 distinct clinical, laboratory, and nursing variables, which has been shown to have prognostic value in some surgical and critical care studies, although its utility in COVID-19 is presently unknown.^30-32^ We hypothesized that the RI might be a useful tool for evaluating disease severity in COVID-19, both for clinical care and for the purposes of propensity score matching. In this observational study, we first analyzed a large, multisite cohort of hospitalized COVID-19 patients by multivariable regression analysis and found a novel prognostic role for the admission RI in predicting in-hospital mortality. Then, incorporating the RI and other measures of illness severity, we performed propensity score matching and multivariable regression analyses and observed significant reductions in in-hospital mortality among hospitalized COVID-19 patients treated with intermediate-dose anticoagulation or aspirin.

## METHODS

### Patients, data collection, and variables

Institutional Review Board approval was obtained for this study; an approved Data Use Agreement between institutions permitted analysis. From March through June 2020, our hospital’s Joint Data Analytics Team (JDAT) identified 4150 hospital encounters in the Yale New Haven Health System with a diagnosis of COVID-19 established via a nasopharyngeal polymerase chain reaction test (Supplementary Table 1). Patients were excluded if they were < 18 years of age (N = 35), had multiple inpatient hospital encounters due to transfer between hospitals or readmission (N = 1247; all such encounters were excluded in these cases), or had missing data (N = 83), yielding an overall study cohort size of 2785 unique patients.

Demographic, clinical, and laboratory data were extracted from each patient’s medical record by JDAT. Established population health registries were used to identify patients with diabetes, hypertension, coronary artery disease (CAD), and congestive heart failure (CHF) (Supplementary Table 2). Inclusion into a population health registry required an encounter diagnosis of the referenced disease state and at least a single, non-abstract patient encounter within the health system in the preceding three years; in addition, either the patient problem list was required to contain the referenced diagnosis, or the patient had to have a minimum of two face-to-face encounters within the previous 12 months. For disease states without established population health registries, ICD-10 codes were used. We defined cardiovascular disease as any of the following: hypertension, diabetes, CAD, myocardial infarction, CHF, atrial fibrillation, stroke, or transient ischemic attack. We categorized body mass index (BMI) according to the U.S. Centers for Disease Control definitions. We categorized the first RI on admission into four quartiles (quartile 1, RI -33 to 43; quartile 2, RI 42 to 65; quartile 3, RI 66 to 79; quartile 4, RI 80 to 99), with the lowest and highest quartiles representing patients with the greatest and least illness severities, respectively.

### Definitions of anticoagulation intensity

For our analysis, each patient was assigned to one anticoagulant group using the following criteria. First, the maximum dose of enoxaparin or heparin received during each patient’s admission was determined. Patients who received a maximum enoxaparin dose of 30-40 mg at a weight-adjusted concentration of < 0.7 mg/kg every 24 hours, enoxaparin 30-40 mg at a weight-adjusted concentration of < 0.4 mg/kg every 12 hours, subcutaneous unfractionated heparin (UFH) 5000 units up to three times per day, or subcutaneous UFH 5000 or 7500 units up to three times per day with a BMI ≥ 40 kg/m2, and who did not receive any other type of documented anticoagulant during their hospitalization, were categorized as prophylactic-dose anticoagulation. Patients who received a maximum enoxaparin dose of ≥ 0.4 and < 0.7 mg/kg every 12 hours or subcutaneous UFH 7500 U at any frequency with a BMI < 40 kg/m2, and who did not receive any other type of anticoagulant during their hospitalization, were categorized as intermediate-dose anticoagulation. Patients who received a maximum enoxaparin dose ≥ 0.7 mg/kg every 12 hours, enoxaparin ≥ 0.7 mg/kg every 24 hours with creatinine clearance < 30 mL/min, enoxaparin ≥ 1.4 mg/kg every 24 hours, intravenous UFH, or intravenous bivalirudin were categorized as therapeutic-dose anticoagulation. Patients who received any other dose of enoxaparin and who did not receive a direct oral anticoagulant (DOAC) or any other therapeutic-dose anticoagulant were categorized as “Alternative enoxaparin dose”. Patients who received a DOAC and no other type of therapeutic-dose anticoagulation were categorized as “DOAC”. All other patients were categorized as “No documented anticoagulation”. Manual chart review was performed in cases with ambiguous data regarding anticoagulation dosing.

### Statistical analyses

The primary outcome in this study was in-hospital death, measured as cumulative incidence of in-hospital death, with cumulative incidence of hospital discharge as a competing risk. Univariable and multivariable regression modeling of subdistribution hazard functions for the primary outcome was performed in all cohorts; we also reported hazard ratios (HR) from competing risks regression.^33^ Variables incorporated into the modeling included demographic factors, medical history, and clinical and laboratory features reflecting disease severity. Propensity score matching was performed on the different cohorts to achieve balance in covariates between patients treated with intermediate-versus prophylactic-dose anticoagulation in the anticoagulation cohort, and separately between patients treated with in-hospital aspirin versus no antiplatelet therapy in the aspirin cohort. Cumulative incidence curves were estimated for nonparametric visualization of in-hospital death and discharge events and tested using Gray’s test in the propensity score-matched anticoagulation and aspirin cohorts^34^; for clarity, only curves for in-hospital death are displayed in the figures.

Propensity scores were calculated within each cohort using multivariable logistic regression models. Propensity scores included covariates that may affect both the likelihood of patients to receive the treatment of interest and the outcome of interest, and that were unbalanced between treatment groups before matching. These variables included a number of patient characteristics as well as markers of disease severity. Matching based on propensity scores incorporating different sets of covariates was performed using a 1:1 nearest-neighbor algorithm, either with a caliper width of 0.25 (anticoagulation cohort) or without a caliper (aspirin cohort). In each analysis, the approach that yielded the best-matched cohort was identified based on the most balanced distribution of propensity scores and the best balance in individual covariates between the two treatment groups.

## RESULTS

In March 2020, our hospital system established antithrombotic guidelines for management of hospitalized COVID-19 patients (Table 1). These guidelines recommended empiric prophylactic- or intermediate-dose anticoagulation in all hospitalized COVID-19 patients based on their D-dimer level, which was measured once or twice daily throughout each patient’s hospital admission; or therapeutic-dose anticoagulation based on clinical suspicion for VTE. Ultimate decisions about anticoagulation dosing were left to the discretion of providers based on their assessments of individual patients.

**Table 1.** Institutional antithrombotic guidelines. Prior to April 3, 2020, all patients were recommended for prophylactic-dose or intermediate-dose anticoagulation, except for those with suspected or radiologically confirmed venous thromboembolism, who were recommended for therapeutic-dose anticoagulation. Prior to April 13, 2020, patients with D-dimer ≥ 10 mg/L fibrinogen equivalent units were recommended for intermediate-dose anticoagulation. On April 13, 2020, the D-dimer threshold for intermediate-dose anticoagulation was changed to 5 mg/L fibrinogen equivalent units. Starting on May 18, 2020, aspirin 81 mg was recommended for all patients. Abbreviations: BID, twice daily; BMI, body mass index; CrCl, creatinine clearance; DOAC, direct oral anticoagulant; FEU, fibrinogen equivalent units; GTT, infusional drip; QD, daily; SC, subcutaneous; TID, three times a day; UFH, unfractionated heparin.

| D-dimer range | Anticoagulation intensity | BMI < 40 kg/m<sup>2</sup> |  | BMI ≥ 40 kg/m<sup>2</sup> |  |
| --- | --- | --- | --- | --- | --- |
| < 5 mg/L FEU | Prophylactic | CrCl, mL/min |  | CrCl, mL/min |  |
|  |  | < 30 | ≥ 30 | < 30 | ≥ 30 |
|  |  | Enoxaparin<br>30 mg SC<br>QD<br>UFH<br>5000 U SC<br>BID-TID | Enoxaparin<br>40 mg SC<br>QD | Enoxaparin<br>40 mg SC<br>QD<br>UFH<br>7500 U SC<br>BID-TID | Enoxaparin<br>40 mg SC<br>BID |
| Prior to April 13, 2020:<br>≥ 10 mg/L FEU<br>April 13, 2020, and<br>after:<br>≥ 5 mg/L FEU | Intermediate | Enoxaparin<br>0.5 mg/kg SC<br>BID<br>UFH<br>7500 U SC<br>BID-TID | Enoxaparin<br>0.5 mg/kg SC<br>BID | Enoxaparin<br>0.5 mg/kg SC<br>BID | Enoxaparin<br>0.5 mg/kg SC<br>BID |
| Suspected or<br>radiologically<br>confirmed VTE | Therapeutic | Enoxaparin<br>1 mg/kg SC<br>QD<br>DOAC<br>UFH GTT | Enoxaparin<br>1 mg/kg SC<br>BID<br>DOAC | Enoxaparin<br>1 mg/kg SC<br>QD<br>DOAC<br>UFH GTT | Enoxaparin<br>1 mg/kg SC<br>BID<br>DOAC |
| May 18, 2020, and after:<br>Aspirin 81 mg (all patients) |  |  |  |  |  |

In the initial version of the guidelines, hospitalized COVID-19 patients with D-dimer < 10 mg/L fibrinogen equivalent units (FEU) were recommended for prophylactic-dose anticoagulation, while patients with D-dimer > 10 mg/L FEU were recommended for intermediate-dose anticoagulation. On April 13, 2020, following discussions with other peer institutions about their anticoagulation practices, the D-dimer threshold for intermediate-dose anticoagulation in our health system’s guidelines was decreased to 5 mg/L FEU.

Early in the pandemic, studies performed at our institution and others supported a role for endotheliopathy and platelet activation in the development of severe COVID-19.^35,36^ Based on this, during the early phases of the pandemic, a number of providers at our institution routinely administered aspirin to COVID-19 patients who were critically ill. On May 18, 2020, aspirin 81 mg daily was added to our hospital system’s treatment guidelines as a recommendation for all hospitalized patients regardless of critical illness. The overall study cohort consisted of 2785 patients (Supplementary Table 3). Half of patients were male (50.1%; N = 1396). The majority were over 60 years old (58.4%; N = 1627). Among all patients, 13.8% (N = 383) died in the hospital; 83.7% (N = 2330) were discharged alive, while 2.6% (N = 72) remained in the hospital at the time of data abstraction. We sought to identify variables significantly associated with disease severity in COVID-19 for use in propensity score matching. To achieve this, we performed multivariable analyses of the overall study cohort, examining associations of in-hospital death with different variables (Table 2). We observed a novel association of low admission RI quartile with increased cumulative incidence of in-hospital death in a model accounting for the competing risk of hospital discharge. Age > 60, male sex, obesity, and the maximum D-dimer level during hospitalization (DDmax) were also significantly associated with in-hospital death, in keeping with prior studies.^37,38^

**Table 2.** Multivariable analysis of in-hospital death in the overall study cohort. Multivariable regression analysis was performed within the overall study cohort to examine the association of in-hospital death with covariates. Cumulative incidence of in-hospital death was evaluated in a competing risks model with hospital discharge, and hazard ratios (HR) for in-hospital death were reported. For the maximum D-dimer level during hospitalization (DDmax), the hazard ratio represents the effect of an increase of one fibrinogen equivalent unit. Abbreviations: CI, 95% confidence interval; DDmax, maximum D-dimer level during hospitalization; HR, hazard ratio; RI, Rothman Index.

|  |  | Cumulative incidence of in-hospital death<br>(competing risks model) |  |  |
| --- | --- | --- | --- | --- |
|  |  | HR for death | CI | P value |
| Age > 60 years |  | 3.545 | 2.599-4.836 | < 0.001 |
| Male sex |  | 1.315 | 1.070-1.618 | 0.009 |
| Obesity |  | 1.356 | 1.101-1.670 | 0.004 |
| Cardiovascular disease |  | 1.014 | 0.799-1.286 | 0.91 |
| African-American |  | 0.850 | 0.670-1.077 | 0.18 |
| DDmax |  | 1.040 | 1.030-1.051 | < 0.001 |
| RI on admission | Quartile 1 | 6.713 | 4.860-9.274 | < 0.001 |
|  | Quartile 2 | 2.764 | 1.958-3.903 | < 0.001 |

### Intermediate-versus prophylactic-dose anticoagulation

To study the potential impact of intermediate-versus prophylactic-dose anticoagulation, we created the “anticoagulation cohort”, a nested cohort of patients from the overall study cohort who were anticoagulated with either a maximum of prophylactic-dose enoxaparin or unfractionated heparin, or a maximum of intermediate-dose enoxaparin or unfractionated heparin (N = 1624). We then performed propensity score matching on patients in the anticoagulation cohort using a number of variables, including age, body mass index (BMI), DDmax, admission RI score, male sex, and African-American race (Supplementary Figure 1). Among all the different combinations of variables tested in patients in the anticoagulation cohort, propensity score matching with age, BMI, DDmax, admission RI score, and African-American race achieved the most balanced distribution of covariates between patients treated with prophylactic-compared to intermediate-dose anticoagulation (Supplementary Table 4).

The final propensity score-matched group of 382 patients from the anticoagulation cohort was well-balanced between patients who received prophylactic-versus intermediate-dose anticoagulation with respect to all variables analyzed except for DDmax, which was higher in patients who received intermediate-dose anticoagulation, reflecting our hospital’s treatment guidelines (Supplementary Table 4). Using this group of propensity score-matched patients, we fit a competing risks multivariable regression model adjusting for age, aspirin and antiplatelet therapy use, male sex, obesity, cardiovascular disease, African-American race, DDmax, and admission RI. Treatment with intermediate-compared to prophylactic-dose anticoagulation was associated with a significantly lower cumulative incidence of in-hospital death on multivariable regression (HR 0.518 [0.308-0.872] (Table 3). Cumulative incidence curves also showed a significant reduction in in-hospital death among propensity score-matched patients in the anticoagulation cohort who were treated with intermediate-compared to prophylactic-dose anticoagulation (Figure 1).

**Table 3.** Multivariable analysis of in-hospital death in the propensity-score matched anticoagulation cohort. Multivariable regression analysis was performed among propensity score-matched patients within the anticoagulation cohort to examine the association of in-hospital death with covariates. Cumulative incidence of in-hospital death was evaluated in a competing risks model with hospital discharge, and hazard ratios (HR) for in-hospital death were reported. For the maximum D-dimer level during hospitalization (DDmax), the hazard ratio represents the effect of an increase of one fibrinogen equivalent unit. Abbreviations: CI, 95% confidence interval; DDmax, maximum D-dimer level during hospitalization; HR, hazard ratio; RI, Rothman Index.

|  |  | Cumulative incidence of in-hospital death |  |  |
| --- | --- | --- | --- | --- |
|  |  | HR for death | CI | P value |
| Intermediate-dose anticoagulation (compared to prophylactic-dose) |  | 0.518 | 0.308-0.872 | 0.013 |
| In-hospital aspirin |  | 0.311 | 0.153-0.634 | 0.001 |
| Home antiplatelet agent use prior to hospitalization |  | 2.663 | 1.335-5.313 | 0.006 |
| Age > 60 years |  | 3.269 | 1.694-6.310 | < 0.001 |
| Male sex |  | 2.255 | 1.283-3.963 | 0.005 |
| Obesity |  | 2.096 | 1.217-3.608 | 0.008 |
| Cardiovascular disease |  | 1.588 | 0.886-2.846 | 0.12 |
| African-American |  | 0.674 | 0.392-1.160 | 0.15 |
| DDmax |  | 1.050 | 1.021-1.080 | < 0.001 |
| RI on admission | Quartile 1 | 10.842 | 4.148-28.341 | < 0.001 |
|  | Quartile 2 | 6.518 | 2.394-17.751 | < 0.001 |

**Figure 1.**
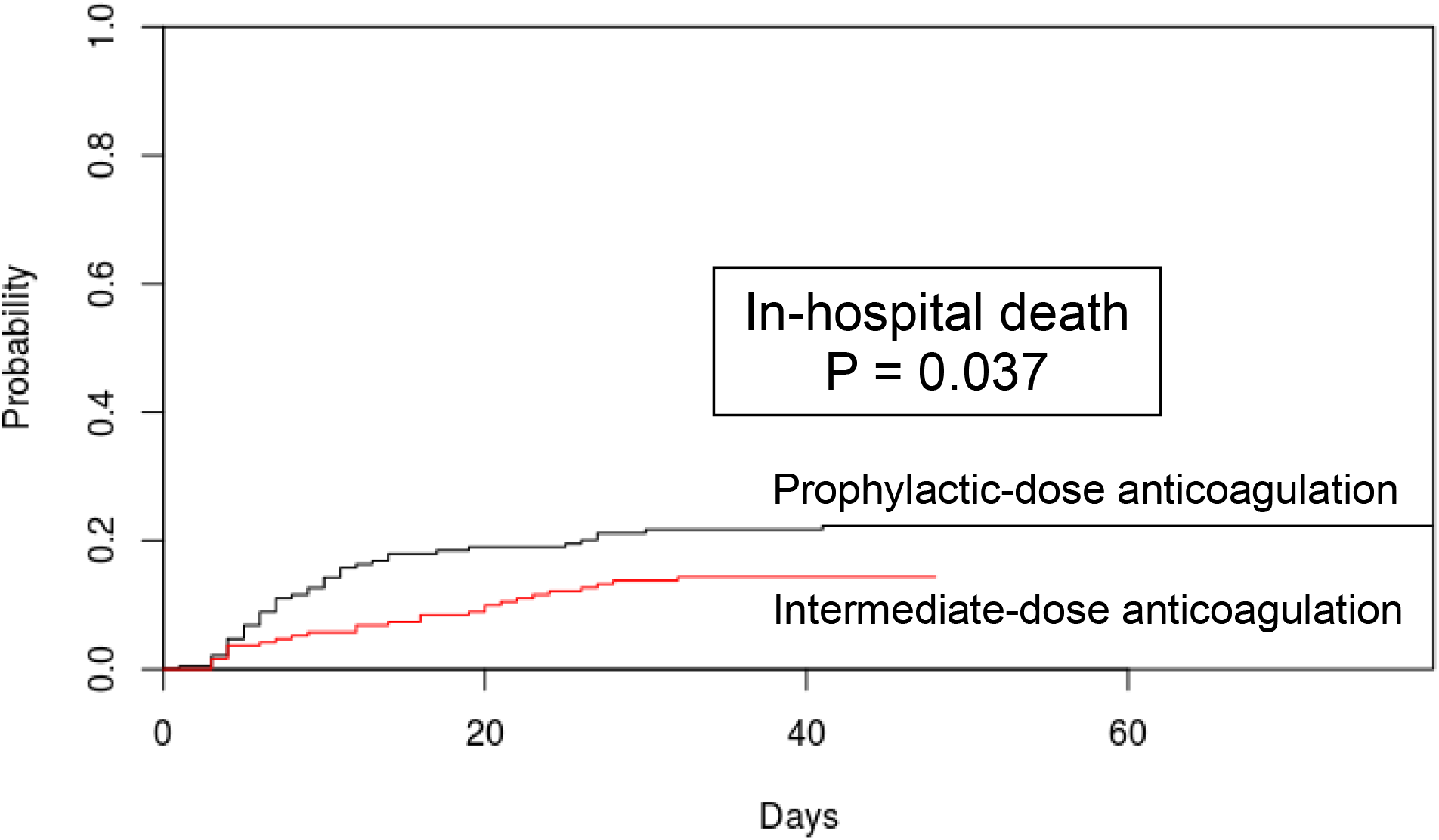
Cumulative incidence of in-hospital death among propensity score-matched patients in the anticoagulation cohort, comparing intermediate-versus prophylactic-dose anticoagulation. Patients were propensity score matched for age, maximum D-dimer level, admission Rothman Index score, body mass index, and African-American race using a random number seed and a caliper width of 0.25. P values from Gray’s test describe differences in cumulative incidence function between intermediate- and prophylactic-dose anticoagulation groups.

### Aspirin versus no antiplatelet therapy

Next, we explored the effects of in-hospital aspirin use. For this analysis, we established the “aspirin cohort”, a nested cohort of patients from the overall study cohort who were not on home antiplatelet therapy prior to admission and received either aspirin or no antiplatelet therapy during their hospitalization (N = 1956). Within the aspirin cohort, we performed propensity score matching for age, DDmax, admission RI, and male sex, which achieved the most balanced distribution of covariates between patients treated with in-hospital aspirin compared to those who received no antiplatelet therapy (Supplementary Figure 2; Supplementary Table 5).

Using this propensity score-matched group of 638 patients, we fit a competing risks multivariable regresson model adjusting for age, anticoagulation other than prophylactic dose, male sex, obesity, cardiovascular disease, African-American race, DDmax, and admission RI; in addition, we included ICU admission as a covariate based on a tendency of providers at our institution to administer aspirin preferentially to critically ill patients earlier on in the pandemic, before aspirin was added onto our hospital’s treatment guidelines. On multivariable analysis of propensity score-matched patients in the aspirin cohort, the use of in-hospital aspirin was associated with a lower cumulative incidence of in-hospital death (HR 0.522 [0.336-0.812]) (Table 4).

**Table 4.** Multivariable analysis of in-hospital death in the propensity-score matched aspirin cohort. Multivariable regression analysis was performed among propensity score-matched patients within the aspirin cohort to examine the association of in-hospital death with covariates. Cumulative incidence of in-hospital death was evaluated in a competing risks model with hospital discharge, and hazard ratios (HR) for in-hospital death were reported. For the maximum D-dimer level during hospitalization (DDmax), the hazard ratio represents the effect of an increase of one fibrinogen equivalent unit. Abbreviations: CI, 95% confidence interval; DDmax, maximum D-dimer level during hospitalization; DOAC, direct oral anticoagulant; ICU, intensive care unit; RI, Rothman Index.

|  |  | Cumulative incidence of in-hospital death |  |  |
| --- | --- | --- | --- | --- |
|  |  | HR for death | CI | P value |
| In-hospital aspirin |  | 0.522 | 0.336-0.812 | 0.004 |
| Anticoagulation other than prophylactic-dose (includes intermediate, therapeutic, DOAC, or other) |  | 2.034 | 1.016-4.074 | 0.045 |
| ICU |  | 3.207 | 1.691-6.080 | < 0.001 |
| Age > 60 years |  | 3.894 | 2.196-6.904 | < 0.001 |
| Male sex |  | 1.227 | 0.777-1.938 | 0.38 |
| Obesity |  | 1.342 | 0.873-2.063 | 0.18 |
| Cardiovascular disease |  | 1.285 | 0.803-2.056 | 0.3 |
| African-American |  | 0.525 | 0.298-0.926 | 0.026 |
| DDmax |  | 1.022 | 0.998-1.047 | 0.069 |
| RI on admission | Quartile 1 | 3.333 | 1.774-6.264 | < 0.001 |
|  | Quartile 2 | 2.022 | 1.048-3.901 | 0.036 |

Separately, we also analyzed outcomes of patients in the aspirin cohort who were admitted after May 18, the date on which our hospital system’s antithrombotic guidelines added a recommendation to administer aspirin to all hospitalized COVID-19 patients (Table 1). For this analysis, we applied propensity score matching for age, DDmax, and admission RI score, which together yielded the most balanced distribution of covariates between aspirin- and non-aspirin-treated patients admitted after May 18, among the different combination of variables tested (Supplementary Figure 3). The final group of 140 propensity score-matched patients was well-balanced between aspirin- and non-aspirin-treated patients with respect to all variables except BMI (Supplementary Table 6). Using this group of propensity score-matched patients, we then fit a competing risks multivariable regression model adjusting for age, anticoagulation other than prophylactic dose, male sex, obesity, cardiovascular disease, African-American race, DDmax, and admission RI. Once again, among patients admitted after May 18, the use of in-hospital aspirin compared to no antiplatelet therapy was associated with a significantly lower cumulative incidence of in-hospital death on multivariable regression (HR 0.036 [0.002-0.576] (Table 5). Cumulative incidence curves showed a significant reduction in in-hospital death among propensity score-matched patients in the aspirin cohort admitted after May 18 who were treated with in-hospital aspirin compared to those who did not receive any antiplatelet therapy (Figure 2).

**Table 5.** Multivariable analysis of in-hospital death in propensity-score matched patients in the aspirin cohort admitted after May 18, 2020. Multivariable regression analysis was performed among propensity score-matched patients within the aspirin cohort admitted after May 18, 2020, in order to examine the association of in-hospital death with covariates. Cumulative incidence of in-hospital death was evaluated in a competing risks model with hospital discharge, and hazard ratios (HR) for in-hospital death were reported. For the maximum D-dimer level during hospitalization (DDmax), the hazard ratio represents the effect of an increase of one fibrinogen equivalent unit. Abbreviations: CI, 95% confidence interval; DDmax, maximum D-dimer level during hospitalization; DOAC, direct oral anticoagulant; RI, Rothman Index.

|  |  | **Cumulative incidence of in-hospital <u>death</u>** |  |  |
| --- | --- | --- | --- | --- |
|  |  | **HR for death** | **CI** | **P value** |
| **In-hospital aspirin** |  | 0.037 | 0.002-0.576 | 0.018 |
| **Anticoagulation other than prophylactic-dose (includes intermediate, therapeutic, DOAC, or other)** |  | 10.879 | 1.965-60.237 | 0.006 |
| **Age > 60 years** |  | 1.278 | 0.357-4.576 | 0.71 |
| **Male sex** |  | 4.132 | 0.762-22.403 | 0.1 |
| **Obesity** |  | 0.552 | 0.129-2.364 | 0.42 |
| **Cardiovascular disease** |  | 0.936 | 0.227-3.858 | 0.93 |
| **African-American** |  | 1.280 | 0.336-4.870 | 0.72 |
| **DDmax** |  | 0.989 | 0.916-1.068 | 0.78 |
| **RI on admission** | **Quartile 1** | 9.413 | 1.435-61.736 | 0.019 |
|  | **Quartile 2** | 1.159 | 0.184-7.301 | 0.88 |

**Figure 2.**
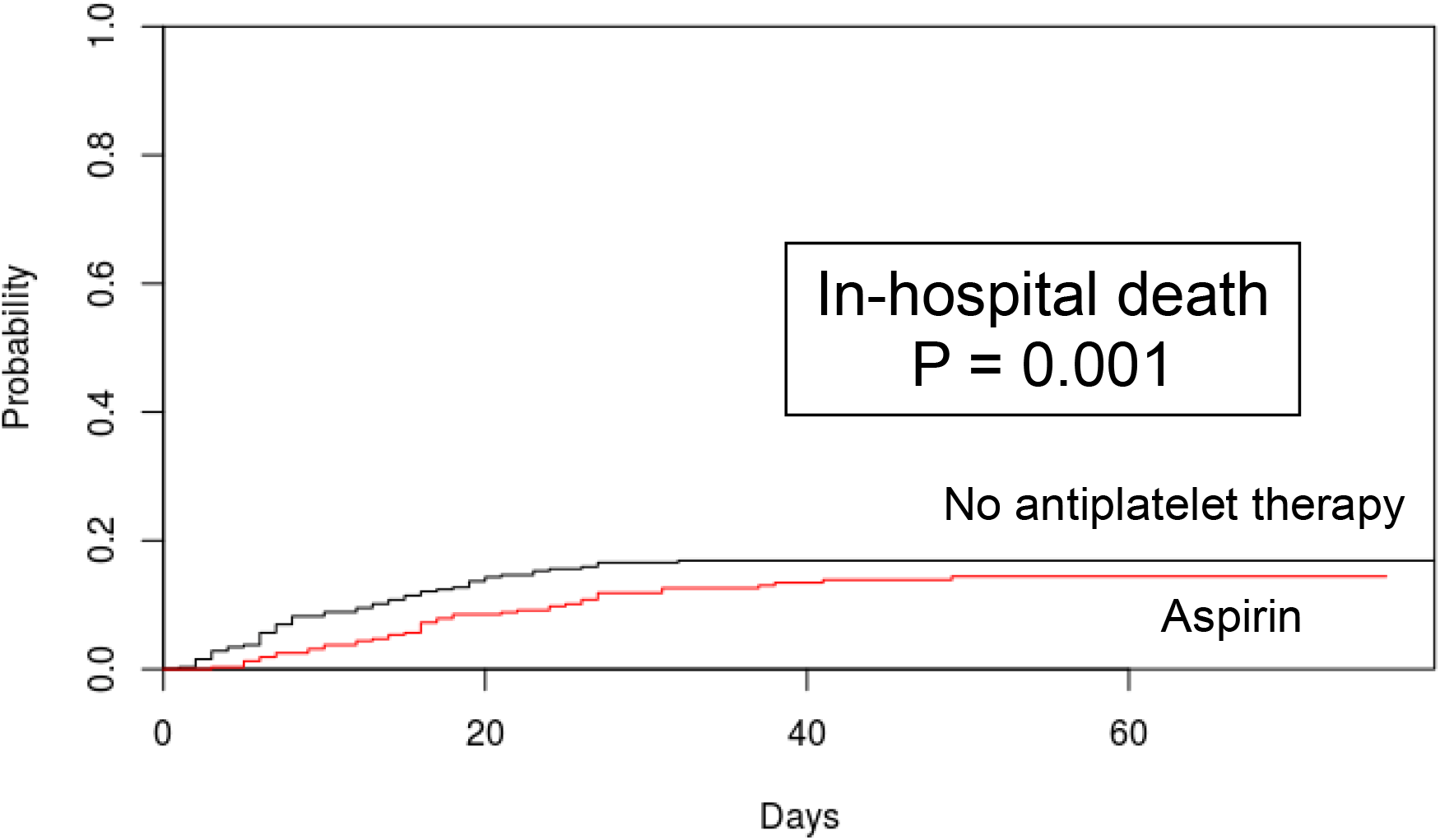
Cumulative incidence of in-hospital death among propensity score-matched patients in the aspirin cohort admitted after May 18, 2020, comparing in-hospital aspirin versus no antiplatelet therapy. Patients were propensity score matched for age, maximum D-dimer level, and admission Rothman Index score. P values from Gray’s test describe differences in cumulative incidence function between patients who received in-hospital aspirin and those who did not.

## DISCUSSION

In our large observational study of hospitalized COVID-19 patients, we report, for the first time, a significantly lower cumulative incidence of in-hospital death among patients who received intermediate-compared to prophylactic-dose anticoagulation, and, separately, among patients who received in-hospital aspirin compared to those who received no antiplatelet therapy. At present, consensus groups differ in their recommendations regarding the use of escalated-intensity anticoagulation in COVID-19 patients who are critically ill, highlighting the uncertainty that exists with this practice.^39-41^ Retrospective studies of therapeutic-compared to prophylactic-dose anticoagulation have reported mixed effects on mortality, while only two small studies have examined outcomes with intermediate-compared to prophylactic-dose anticoagulation, largely focusing on venous thromboembolism rates, again with conflicting results.^7,15,16,18-22,42,43^ More recently, the ACTIV randomized clinical trial showed futility of therapeutic-compared to prophylactic-dose anticoagulation in critically ill COVID-19 patients, leading to a pause in further recruitment of critically ill patients for that trial. Our analysis is the first large-scale study to specifically examine intermediate- and prophylactic-dose anticoagulation in COVID-19. In contrast to many other studies, we utilized propensity score matching and multivariable regression analysis in order to diminish treatment selection bias by generating treatment and control groups with well-balanced covariates, thereby allowing for a more reliable comparison of potential treatment effects.^27^ Our findings suggest that there may be a beneficial role for intermediate-dose anticoagulation in the treatment of hospitalized COVID-19 patients, although we await the results of several randomized controlled trials to definitively address this question.

At present, no consensus guidelines are available regarding aspirin use in COVID-19, reflecting a paucity of data in this regard. A biological rationale to support the use of aspirin in COVID-19 may reside in the treatment of other microvascular thrombotic diseases such as thrombotic thrombocytopenic purpura, where antiplatelet agents may have a role.^44^ Recently, one other retrospective study reported improved in-hospital mortality in COVID-19 patients who received aspirin within one week before or 24 hours after admission.^25^ Despite drawing similar conclusions, our study adopted a more rigorous methodological approach through the use of propensity score matching to account for differences in illness severity among patients, enabling us to more accurately compare different treatment effects. In addition, we excluded patients on home aspirin in order to minimize confounding effects from underlying cardiovascular disease.

Our analysis also reveals a novel role for the admission RI as a prognostic tool for evaluating the risk of in-hospital mortality in COVID-19. The RI, which synthesizes multiple clinical, laboratory, and nursing assessment variables into a single score, has been shown to have predictive value for assessing mortality and readmission rates in some critical care and surgical studies, although its applicability to COVID-19 has not been previously tested.^30-32^ In our hospital system, the RI is calculated automatically by our electronic health record system upon admission and hourly throughout a patient’s hospitalization, rendering it readily accessible to enable its use in real-time clinical decision-making.^32^ Additional studies are warranted to further explore the potential role for the RI in assessing disease severity and guiding clinical interventions in COVID-19.

Our study has several limitations, beyond its retrospective nature. Overall provider adherence to our institution’s COVID-19 treatment guidelines was subject to provider preference, although many of the confounding factors that would have resulted from such bias were accounted for through our use of propensity score matching and multivariable regression analysis. Heterogeneity in the number of doses of intermediate-dose anticoagulation or aspirin that each patient received during their hospitalization may have biased our analysis against the detection of some significant associations by including patients in the intervention group who received limited exposure to the intervention. A possible improvement in clinical outcomes of hospitalized patients with COVID-19 over time could have biased some of our findings, although in our analysis of patients in the aspirin cohort admitted after May 18, a significant reduction in in-hospital mortality with aspirin use was still observed despite the later, shortened timeframe of the specific study population analyzed. We did not examine other COVID-19 therapies that patients may have received and did not examine VTE rates, as only a small percentage of patients in our hospital system underwent VTE-specific imaging in order to limit excess healthcare worker exposure to COVID-19.

In summary, in our large, observational study of hospitalized patients with COVID-19, using propensity score matching and multivariable regression analyses, we observed a mortality benefit with intermediate-compared to prophylactic-dose anticoagulation and, separately, with in-hospital aspirin compared to no antiplatelet therapy. Our findings suggest that increased-intensity anticoagulation and antiplatelet therapy may be beneficial in the treatment of COVID-19. We await the results of several randomized clinical trials to more definitively elucidate the impact of these therapies in COVID-19.

## Data Availability

All authors have access to the data provided in this manuscript.

## Acknowledgements

The authors would like to thank all providers, health care workers, and staff for their tireless dedication to the care of patients with COVID-19. We dedicate this work to all individuals afflicted by the pandemic. An earlier version of this study was presented at an Oral Session at the 62^nd^ Annual Meeting of the American Society of Hematology.

## Author Contributions

G.G., M.L.M., Yiwen Liu, R.F., D.S.N., K.A.O., and A.I.L. designed the study. M.L.M., R.F., K.A., E.C., N.D., C.K., and Yuxin Liu performed chart abstractions. M.M., D.M., S.W., C.P., R.D.B., C.I.O.C., H.J.C., and A.B.P. contributed valuable ideas. M.L.M., G.G., Yiwen Liu, R.F., D.S.N., K.A.O., and A.I.L. wrote the manuscript, and all authors participated in editing the manuscript. M.L.M., G.G., Yiwen Liu, and R.F. contributed equally as first authors. D.S.N., K.A.O., and A.I.L. contributed equally as senior investigators.

## Financial/nonfinancial disclosures

No conflict of interest exists for any author on this manuscript. This work was supported by a gift donation from Jack Levin and a separate anonymous donation to the Benign Hematology program at Yale, the DeLuca Foundation to fund hematology research at Yale, and the National Institutes of Health (grant HL142818 to H.J.C., and GM136651 and HL139116 to M.L.).

## Role of the sponsors

The funder of the study had no role in study design, data collection, data analysis, data interpretation, or writing of the report.

---

**Supplementary Table 1.** Hospitals in the Yale New Haven Health System.

| **Yale New Haven Health System hospitals** |
| --- |
| Yale New Haven Hospital |
| Bridgeport Hospital |
| Greenwich Hospital |
| Lawrence & Memorial Hospital |
| St. Raphael Campus |
| Westerly Hospital |

**Supplementary Table 2.** Registry exclusions.

| Population | Exclusions related to disease diagnosis |
| --- | --- |
| Congestive heart failure | • Hypertensive heart failure<br>• Cor pulmonale<br>• Neonatal cardiovascular disorder<br>• Cardiac complication of procedure<br>• Cardiorenal syndrome<br>• Right heart failure due to pulmonary hypertension<br>• Hypertensive heart disease<br>• Complication of pregnancy, childbirth and/or the puerperium |
| Coronary artery disease | • None |
| Diabetes Mellitus | • Diabetes mellitus during pregnancy, childbirth, and the puerperium<br>• Fetal hypertrophic cardiomyopathy<br>• Gestational diabetes mellitus, controlled<br>• Steroid-induced diabetes |
| Hypertension | • Pregnancy, childbirth and puerperium finding<br>• Complication of pregnancy<br>• Pre-existing hypertension in obstetric context<br>• Chronic hypertension in obstetric context<br>• Essential hypertension in obstetric context<br>• Labile hypertension due to being in a clinical environment<br>• Transient hypertension<br>• Neonatal hypertension<br>• Postoperative hypertension |

**Supplementary Table 3.**
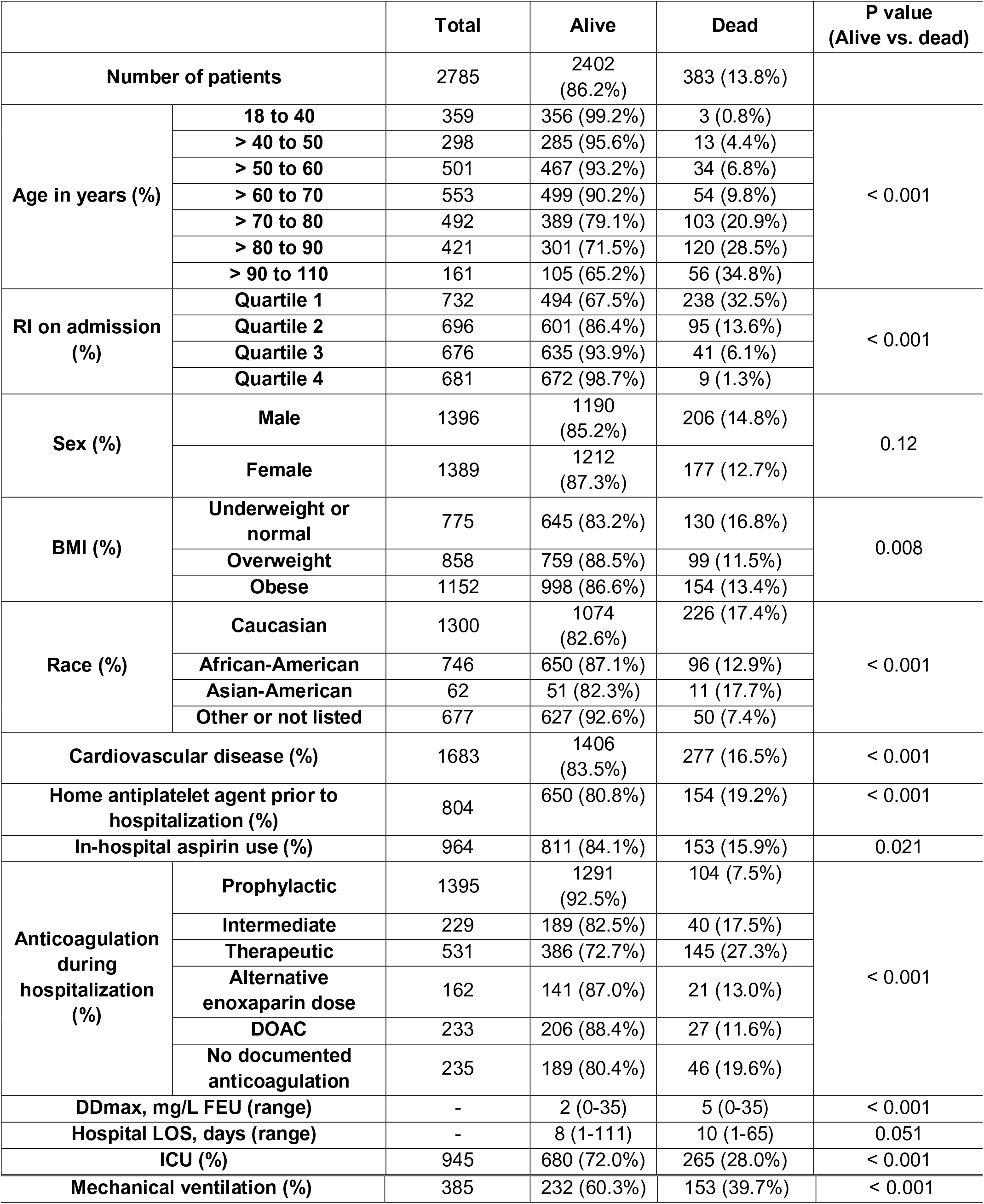
Patient characteristics in overall study cohort. “Anticoagulation during hospitalization” is based upon the maximum intensity of anticoagulation that each patient received during hospitalization, with each patient falling into one category (as described in the “Methods”). “Prophylactic” anticoagulation refers to patients who received a maximum of prophylactic-dose enoxaparin or subcutaneous unfractionated heparin. “Intermediate” anticoagulation refers to patients who received a maximum of intermediate-dose enoxaparin or subcutaneous unfractionated heparin. “Therapeutic” anticoagulation refers to patients who received a maximum of therapeutic-dose enoxaparin, intravenous heparin, or bivalirudin. “Alternative enoxaparin dose” refers to patients who received a maximum dose of enoxaparin that was not able to be categorized as prophylactic, intermediate, or therapeutic. “No documented anticoagulation” refers to patients who did not receive enoxaparin, heparin, bivalirudin, or a direct oral anticoagulant during their hospitalization. Abbreviations: BMI, body mass index; DDmax, maximum D-dimer level during hospitalization; DOAC, direct oral anticoagulant; FEU, fibrinogen equivalent units; ICU, intensive care unit; LOS, length of stay, expressed as median; RI, Rothman Index.

**Supplementary Table 4.** Patient characteristics in the propensity score-matched anticoagulation cohort. Patients were propensity score matched for age, maximum D-dimer level, admission Rothman Index score, body mass index, and African-American race using a random number seed and a caliper width of 0.25. P values describe differences between prophylactic- and intermediate-dose anticoagulation groups. Abbreviations: BMI, body mass index; DDmax, maximum D-dimer level during hospitalization, expressed as median; FEU, fibrinogen equivalent units; RI, Rothman Index.

|  |  | Total | Prophylactic-dose anticoagulation | Intermediate-dose anticoagulation | P value |
| --- | --- | --- | --- | --- | --- |
| **Number of patients** |  | 382 | 191 | 191 | - |
| **Age** | **≤ 60 years** | 148 | 73 | 75 | 0.83 |
|  | **> 60 years** | 234 | 118 | 116 |  |
| **RI on admission** | **Quartile 1** | 145 | 78 | 67 | 0.40 |
|  | **Quartile 2** | 91 | 46 | 45 |  |
|  | **Quartiles 3 and 4** | 146 | 67 | 79 |  |
| **Sex** | **Male** | 195 | 96 | 99 | 0.76 |
|  | **Female** | 187 | 95 | 92 |  |
| **BMI** | **Obese** | 181 | 82 | 99 | 0.082 |
|  | **Other** | 201 | 109 | 92 |  |
| **Race** | **African-American** | 145 | 74 | 71 | 0.75 |
|  | **Other** | 237 | 117 | 120 |  |
| **Cardiovascular disease** |  | 237 | 113 | 124 | 0.25 |
| **Home antiplatelet agent prior to hospitalization** |  | 127 | 63 | 64 | 0.91 |
| **In-hospital aspirin** |  | 149 | 69 | 80 | 0.29 |
| **DDmax, mg/L FEU (range)** |  | - | 4 (0-34) | 6 (0-35) | 0.003 |

**Supplementary Table 5.** Patient characteristics in the propensity score-matched aspirin cohort. Patients were propensity score matched for age, maximum D-dimer level, admission Rothman Index score, and male sex. P values describe differences between in-hospital aspirin and no in-hospital aspirin groups. Abbreviations: BMI, body mass index; DDmax, maximum D-dimer value during first 30 days of hospitalization, expressed as median; DOAC, direct oral anticoagulant; FEU, fibrinogen equivalent units; RI, Rothman Index.

|  |  | Total | No in-hospital aspirin | In-hospital aspirin | P value |
| --- | --- | --- | --- | --- | --- |
| Number of patients |  | 638 | 319 | 319 | - |
| Age | ≤ 60 years | 346 | 146 | 146 | > 0.99 |
|  | > 60 years | 292 | 173 | 173 |  |
| RI on admission | Quartile 1 | 161 | 72 | 89 | 0.18 |
|  | Quartile 2 | 158 | 87 | 71 |  |
|  | Quartiles 3 and 4 | 319 | 160 | 159 |  |
| Sex | Male | 404 | 202 | 202 | > 0.99 |
|  | Female | 234 | 117 | 117 |  |
| BMI | Obese | 269 | 138 | 131 | 0.57 |
|  | Other | 369 | 181 | 188 |  |
| Race | African-American | 149 | 69 | 80 | 0.30 |
|  | Other | 489 | 250 | 239 |  |
| Cardiovascular disease |  | 351 | 174 | 177 | 0.81 |
| Anticoagulation dose group | Prophylactic | 288 | 163 | 125 | 0.003 |
|  | Intermediate, therapeutic, DOAC, or other | 350 | 156 | 194 |  |
| DDmax, mg/L FEU (range) |  | - | 2 (0-35) | 3 (0-34) | 0.18 |

**Supplementary Table 6.**
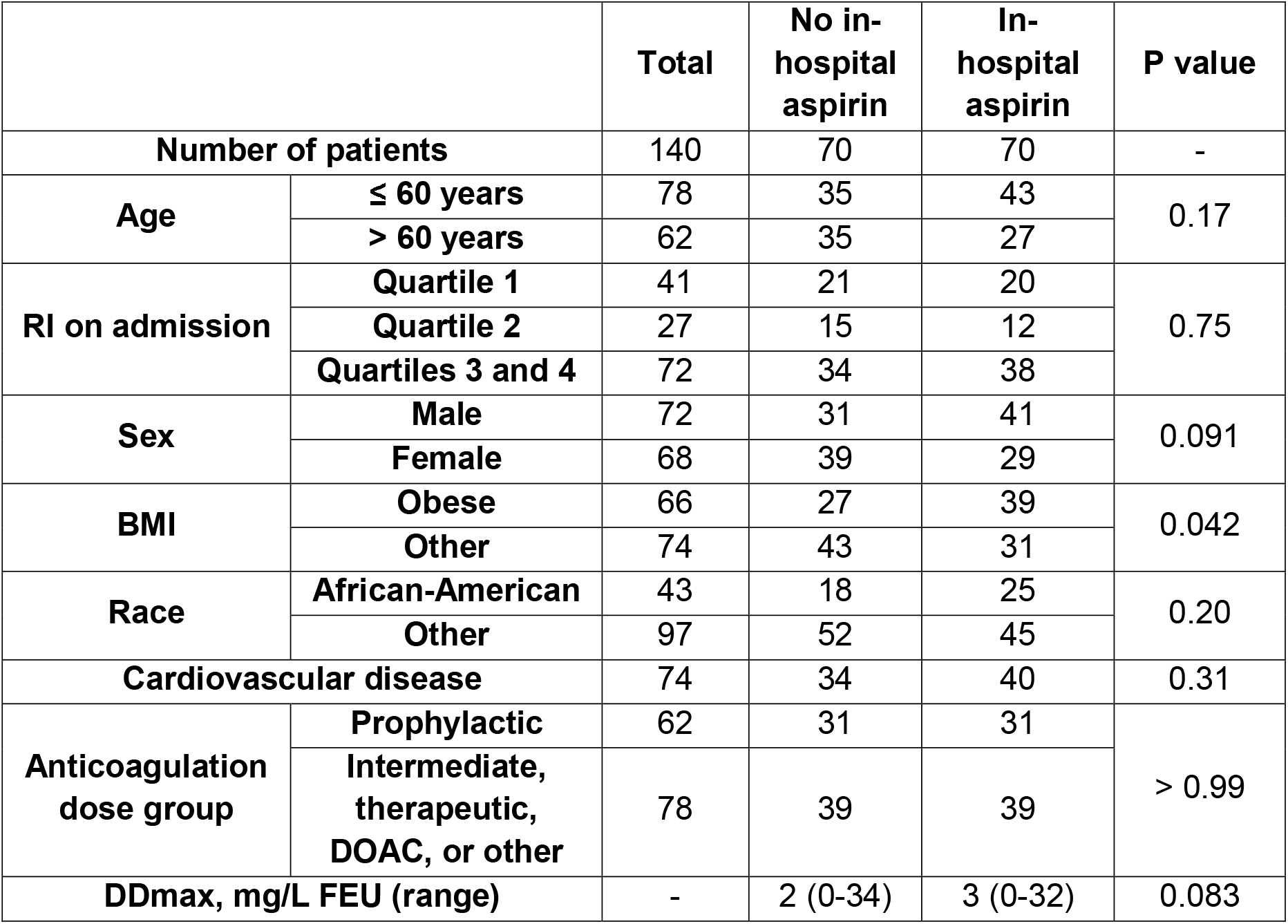
Patient characteristics in the aspirin cohort admitted after May 18, 2020, with propensity score matching. Patients in the aspirin cohort who were admitted after May 18, 2020 were propensity score matched for age, maximum D-dimer level, and admission Rothman Index score. P values describe differences between in-hospital aspirin and no in-hospital aspirin groups. Abbreviations: BMI, body mass index; DDmax, maximum D-dimer level during hospitalization, expressed as median; DOAC, direct oral anticoagulant; FEU, fibrinogen equivalent units; ICU, intensive care unit admission or transfer; RI, Rothman Index.

|  |  | Total | No in-hospital aspirin | In-hospital aspirin | P value |
| --- | --- | --- | --- | --- | --- |
| Number of patients |  | 140 | 70 | 70 | - |
| Age | ≤ 60 years | 78 | 35 | 43 | 0.17 |
|  | > 60 years | 62 | 35 | 27 |  |
| RI on admission | Quartile 1 | 41 | 21 | 20 | 0.75 |
|  | Quartile 2 | 27 | 15 | 12 |  |
|  | Quartiles 3 and 4 | 72 | 34 | 38 |  |
| Sex | Male | 72 | 31 | 41 | 0.091 |
|  | Female | 68 | 39 | 29 |  |
| BMI | Obese | 66 | 27 | 39 | 0.042 |
|  | Other | 74 | 43 | 31 |  |
| Race | African-American | 43 | 18 | 25 | 0.20 |
|  | Other | 97 | 52 | 45 |  |
| Cardiovascular disease |  | 74 | 34 | 40 | 0.31 |
| Anticoagulation dose group | Prophylactic | 62 | 31 | 31 | > 0.99 |
|  | Intermediate, therapeutic, DOAC, or other | 78 | 39 | 39 |  |
| DDmax, mg/L FEU (range) |  | - | 2 (0-34) | 3 (0-32) | 0.083 |

**Supplementary Figure 1.**
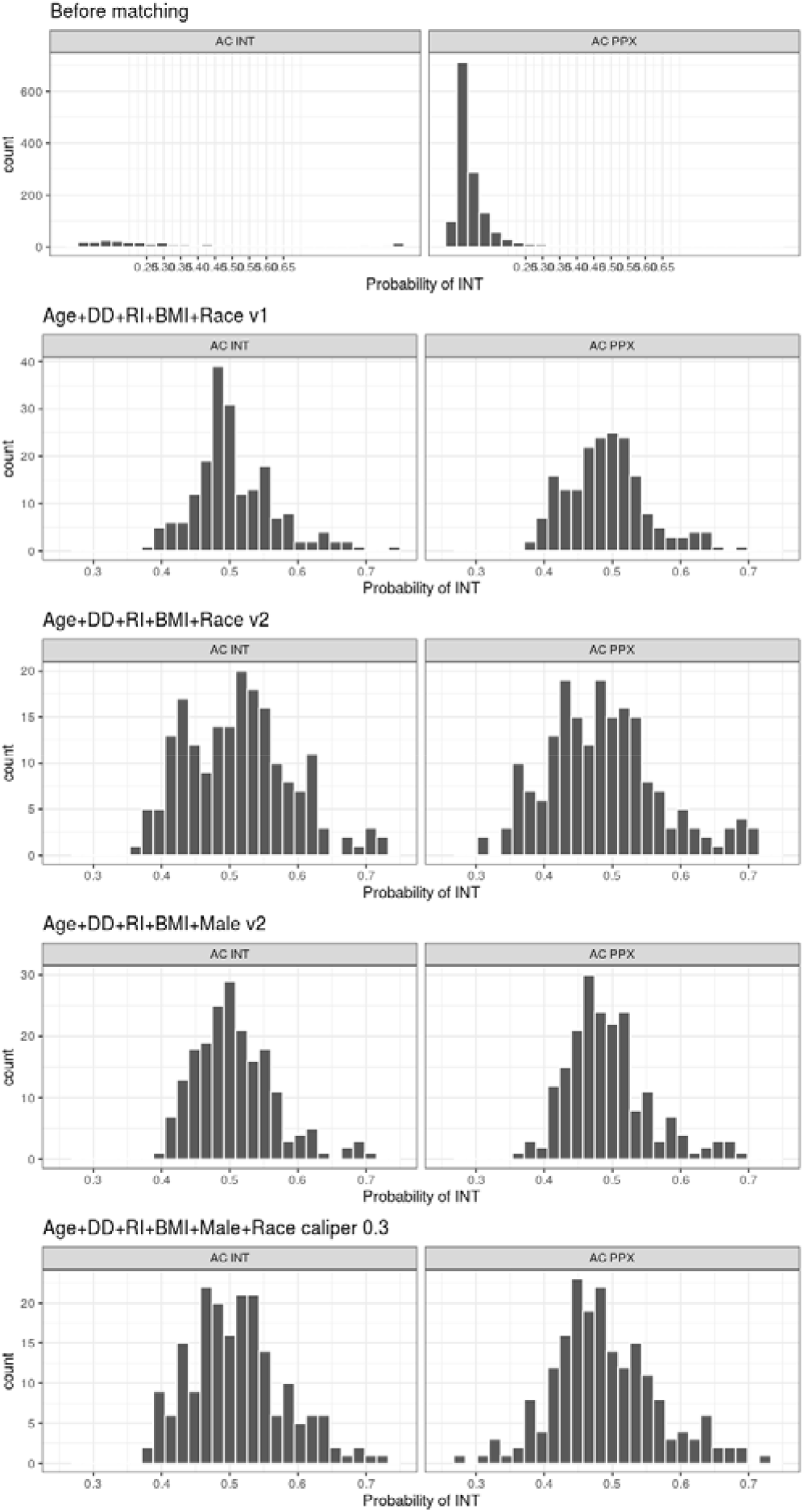
Distribution of propensity scores before and after propensity score matching in the anticoagulation cohort. Propensity scores were calculated using a caliper width of 0.25 and different combinations of variables in the anticoagulation cohort. For each combination of variables, the distribution of propensity scores in patients who received prophylactic- vs. intermediate-dose anticoagulation was compared. Among patients in the anticoagulation cohort, the best-matched scores incorporated age, maximum D-dimer, admission Rothman Index, body mass index, and race with a random number generator seed of 3000 (“Age + DD + RI + BMI + Race v1”). Key: “DD”, maximum D-dimer level during hospitalization; “Male”, male sex; “Race”, African-American race; “Probability of INT”, probability of receiving intermediate-dose anticoagulation; “RI”, admission Rothman Index; “v1”, random number generator seed 3000; “v2”, random number generator seed 1000.

**Supplementary Figure 2.**
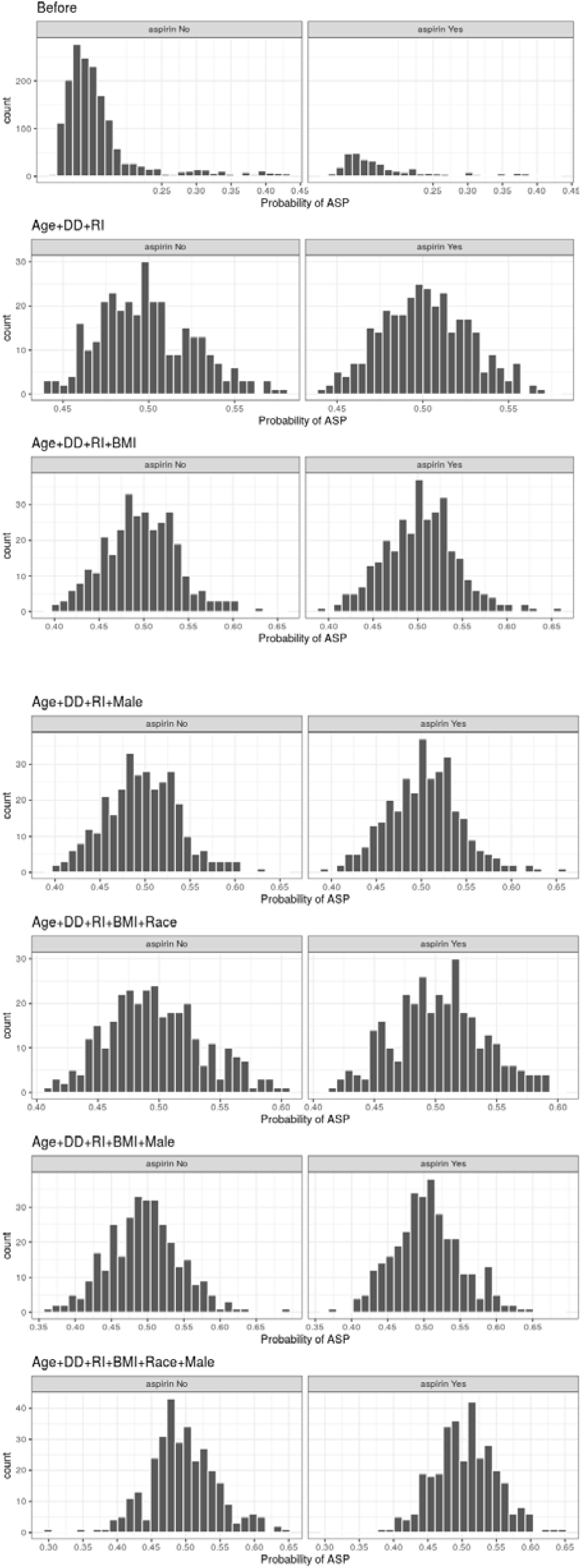
Distribution of propensity scores before and after propensity score matching in the aspirin cohort. Propensity scores were calculated using different combinations of variables in the aspirin cohort. For each combination of variables, the distribution of propensity scores between patients who received in-hospital aspirin versus no antiplatelet therapy was compared. Among patients in the aspirin cohort, the best-matched scores incorporated age, maximum D-dimer, admission Rothman Index, and male sex (“Age + DD + RI + Male”). Key: “DD”, maximum D-dimer level during hospitalization; “Male”, male sex; “Race”, African-American race; “Probability of ASP”, probability of receiving in-hospital aspirin; “RI”, admission Rothman Index.

**Supplementary Figure 3.**
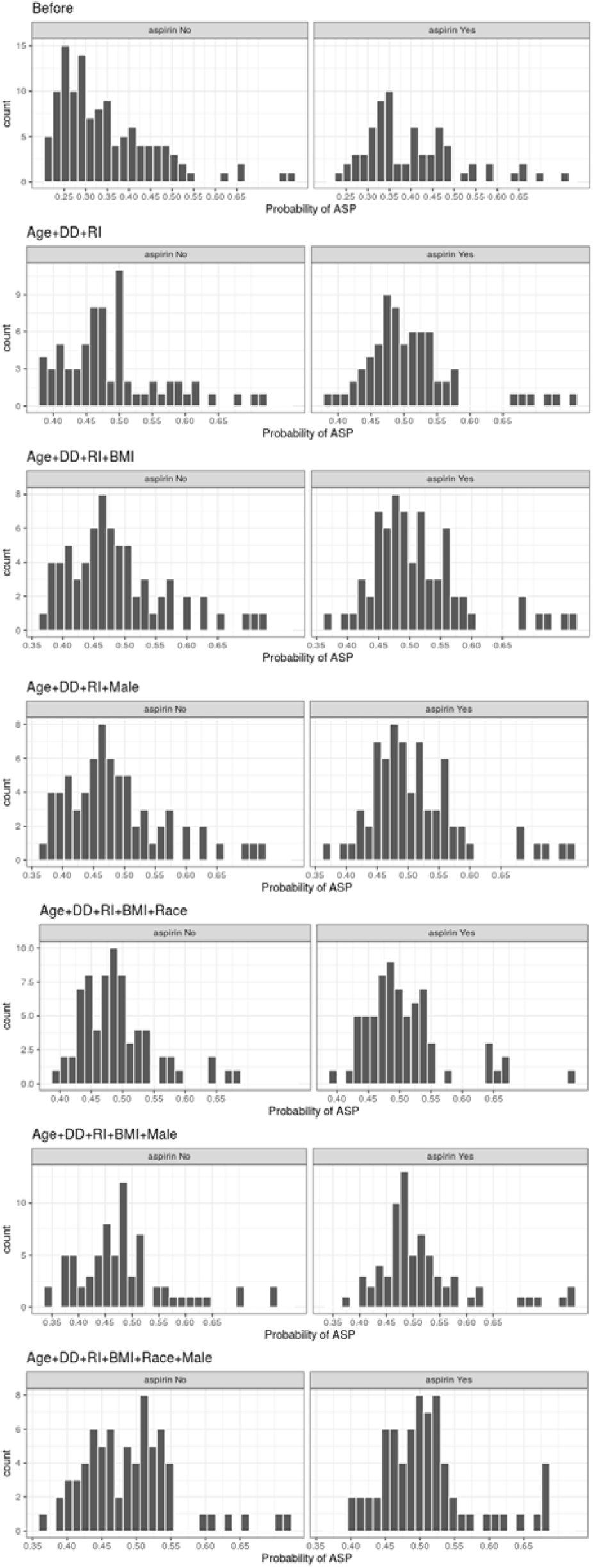
Distribution of propensity scores before and after propensity score matching in the aspirin cohort for patients admitted after May 18, 2020. Propensity scores were calculated using different combinations of variables in the aspirin cohort admitted after May 18. For each combination of variables, the distribution of propensity scores between patients who received in-hospital aspirin versus no antiplatelet therapy was compared. Among patients in the aspirin cohort admitted after May 18, 2020, the best-matched scores incorporated age, maximum D-dimer, and admission Rothman Index (“Age + DD + RI”). Key: “DD”, maximum D-dimer level during hospitalization; “Male”, male sex; “Race”, African-American race; “Probability of ASP”, probability of receiving in-hospital aspirin; “RI”, admission Rothman Index.

## Notes

### Competing Interest Statement

The authors have declared no competing interest.

### Author Declarations

This study was approved by the Yale Institutional Review Board (HIC 2000027792). An additional, approved Data Use Agreement between Yale and Dana-Farber Cancer Institute permitted analysis.

## REFERENCES

1. Al-Samkari H, Karp Leaf RS, Dzik WH, et al. COVID-19 and coagulation: bleeding and thrombotic manifestations of SARS-CoV-2 infection. Blood 2020;136:489–500.

2. Kashi M, Jacquin A, Dakhil B, et al. Severe arterial thrombosis associated with Covid-19 infection. Thromb Res 2020;192:75–7.

3. Merkler AE, Parikh NS, Mir S, et al. Risk of Ischemic Stroke in Patients With Coronavirus Disease 2019 (COVID-19) vs Patients With Influenza. JAMA Neurol 2020.

4. Carsana L, Sonzogni A, Nasr A, et al. Pulmonary post-mortem findings in a series of COVID-19 cases from northern Italy: a two-centre descriptive study. Lancet Infect Dis 2020;20:1135–40.

5. Ackermann M, Verleden SE, Kuehnel M, et al. Pulmonary Vascular Endothelialitis, Thrombosis, and Angiogenesis in Covid-19. N Engl J Med 2020;383:120–8.

6. Nopp S, Moik F, Jilma B, Pabinger I, Ay C. Risk of venous thromboembolism in patients with COVID-19: A systematic review and meta-analysis. Res Pract Thromb Haemost 2020.

7. Trigonis RA, Holt DB, Yuan R, et al. Incidence of Venous Thromboembolism in Critically Ill Coronavirus Disease 2019 Patients Receiving Prophylactic Anticoagulation. Crit Care Med 2020;48:e805–e8.

8. Petrilli CM, Jones SA, Yang J, et al. Factors associated with hospital admission and critical illness among 5279 people with coronavirus disease 2019 in New York City: prospective cohort study. BMJ 2020;369:m1966.

9. Zhou F, Yu T, Du R, et al. Clinical course and risk factors for mortality of adult inpatients with COVID-19 in Wuhan, China: a retrospective cohort study. Lancet 2020;395:1054–62.

10. Barrett CD, Moore HB, Yaffe MB, Moore EE. ISTH interim guidance on recognition and management of coagulopathy in COVID-19: A comment. J Thromb Haemost 2020;18:2060–3.

11. Connors JM, Levy JH. COVID-19 and its implications for thrombosis and anticoagulation. Blood 2020;135:2033–40.

12. Ferrandis R, Llau JV, Quintana M, et al. COVID-19: opening a new paradigm in thromboprophylaxis for critically ill patients? Crit Care 2020;24:332.

13. Adam EH, Zacharowski K, Miesbach W. A comprehensive assessment of the coagulation profile in critically ill COVID-19 patients. Thromb Res 2020;194:42–4.

14. Chowdhury JF, Moores LK, Connors JM. Anticoagulation in Hospitalized Patients with Covid-19. N Engl J Med 2020;383:1675–8.

15. Nadkarni GN, Lala A, Bagiella E, et al. Anticoagulation, Bleeding, Mortality, and Pathology in Hospitalized Patients With COVID-19. J Am Coll Cardiol 2020;76:1815–26.

16. Paranjpe I, Fuster V, Lala A, et al. Association of Treatment Dose Anticoagulation With In-Hospital Survival Among Hospitalized Patients With COVID-19. J Am Coll Cardiol 2020;76:122–4.

17. Tang N, Li D, Wang X, Sun Z. Abnormal coagulation parameters are associated with poor prognosis in patients with novel coronavirus pneumonia. Journal of Thrombosis and Haemostasis 2020.

18. Ferguson J, Volk S, Vondracek T, Flanigan J, Chernaik A. Empiric Therapeutic Anticoagulation and Mortality in Critically Ill Patients With Respiratory Failure From SARS-CoV- 2: A Retrospective Cohort Study. J Clin Pharmacol 2020;60:1411–5.

19. Pesavento R, Ceccato D, Pasquetto G, et al. The hazard of (sub)therapeutic doses of anticoagulants in non-critically ill patients with Covid-19: The Padua province experience. J Thromb Haemost 2020.

20. Ionescu FJ, I.; Nair, G. B.; Konde, A. S.; Petrescu, I.; Anusim, N.; Jindal, V.; Gaikazian, S.; Anderson, J.; Huben, M. T.; Stender, M.; Zimmer, M. S. Increasing doses of anticoagulation are associated with improved survival in hospitalized COVID-19 patients [abstract]. Blood 2020;136:22.

21. Al-Samkari HG, S.; Karp Leaf, R.; Wang, W.; Rosovsky, R.; Bauer, K.; Leaf, D.; STOP- COVID Investigators. Thrombosis, bleeding, and the effect of anticoagulation on survival in critically ill patients with COVID-19 in the United States [abstract]. Res Pract Thromb Haemost 2020;4.

22. Ho GD, J. R.; Schmittdiel, J.; Kavecansky, J.; Tavakoli, J.; Pai, A.. Anticoagulant and antiplatelet use not associated with improvement in severe outcomes in COVID-19 patients [abstract]. Blood 2020;136:59.

23. Nakazawa D, Ishizu A. Immunothrombosis in severe COVID-19. EBioMedicine 2020;59:102942.

24. Lowenstein CJ, Solomon SD. Severe COVID-19 Is a Microvascular Disease. Circulation 2020;142:1609–11.

25. Chow JH, Khanna AK, Kethireddy S, et al. Aspirin Use is Associated with Decreased Mechanical Ventilation, ICU Admission, and In-Hospital Mortality in Hospitalized Patients with COVID-19. Anesth Analg 2020.

26. Hernan MA, Alonso A, Logan R, et al. Observational studies analyzed like randomized experiments: an application to postmenopausal hormone therapy and coronary heart disease. Epidemiology 2008;19:766–79.

27. Sturmer T, Wyss R, Glynn RJ, Brookhart MA. Propensity scores for confounder adjustment when assessing the effects of medical interventions using nonexperimental study designs. J Intern Med 2014;275:570–80.

28. Geleris J, Sun Y, Platt J, et al. Observational Study of Hydroxychloroquine in Hospitalized Patients with Covid-19. N Engl J Med 2020;382:2411–8.

29. Reynolds HR, Adhikari S, Pulgarin C, et al. Renin-Angiotensin-Aldosterone System Inhibitors and Risk of Covid-19. N Engl J Med 2020;382:2441–8.

30. Alarhayem AQ, Muir MT, Jenkins DJ, et al. Application of electronic medical record-derived analytics in critical care: Rothman Index predicts mortality and readmissions in surgical intensive care unit patients. J Trauma Acute Care Surg 2019;86:635–41.

31. Wengerter BC, Pei KY, Asuzu D, Davis KA. Rothman Index variability predicts clinical deterioration and rapid response activation. Am J Surg 2018;215:37–41.

32. Rothman MJ, Rothman SI, Beals Jt. Development and validation of a continuous measure of patient condition using the Electronic Medical Record. J Biomed Inform 2013;46:837–48.

33. Fine JP, Gray RJ. A Proportional Hazards Model for the Subdistribution of a Competing Risk. J Amer Statist Assoc 1999;94:496–509.

34. Gray RJ. A Class of K-Sample Tests for Comparing the Cumulative Incidence of a Competing Risk. Ann Statistics 1988;16:1141–54.

35. Goshua G, Pine AB, Meizlish ML, et al. Endotheliopathy in COVID-19-associated coagulopathy: evidence from a single-centre, cross-sectional study. Lancet Haematol 2020;7:e575–e82.

36. Manne BK, Denorme F, Middleton EA, et al. Platelet gene expression and function in patients with COVID-19. Blood 2020;136:1317–29.

37. Gupta S, Hayek SS, Wang W, et al. Factors Associated With Death in Critically Ill Patients With Coronavirus Disease 2019 in the US. JAMA Intern Med 2020.

38. Zhang L, Yan X, Fan Q, et al. D-dimer levels on admission to predict in-hospital mortality in patients with Covid-19. J Thromb Haemost 2020;18:1324–9.

39. Barnes GD, Burnett A, Allen A, et al. Thromboembolism and anticoagulant therapy during the COVID-19 pandemic: interim clinical guidance from the anticoagulation forum. J Thromb Thrombolysis 2020;50:72–81.

40. Moores LK, Tritschler T, Brosnahan S, et al. Prevention, Diagnosis, and Treatment of VTE in Patients With Coronavirus Disease 2019: CHEST Guideline and Expert Panel Report. Chest 2020;158:1143–63.

41. Spyropoulos AC, Levy JH, Ageno W, et al. Scientific and Standardization Committee communication: Clinical guidance on the diagnosis, prevention, and treatment of venous thromboembolism in hospitalized patients with COVID-19. J Thromb Haemost 2020;18:1859–65.

42. Tang N, Bai H, Chen X, Gong J, Li D, Sun Z. Anticoagulant treatment is associated with decreased mortality in severe coronavirus disease 2019 patients with coagulopathy. J Thromb Haemost 2020;18:1094–9.

43. Taccone FS, Gevenois PA, Peluso L, et al. Higher Intensity Thromboprophylaxis Regimens and Pulmonary Embolism in Critically Ill Coronavirus Disease 2019 Patients. Crit Care Med 2020;48:e1087–e90.

44. Blombery P, Scully M. Management of thrombotic thrombocytopenic purpura: current perspectives. J Blood Med 2014;5:15–23.

